# DBToken: A Database Tokenizer for Medical Event Foundation Models

**DOI:** 10.64898/2026.08.18.26360487

**Authors:** Ikgyu Shin, Kent McCann, Giacomo Marino, Uzair Tahamid Siam, Huan Li, Erica Stutz, Ruthvik Edara, Andrew J Loza

**Author notes:** **Corresponding author:** Andrew J. Loza, MD, PhD, Department of Biomedical Informatics and Data Science, Yale School of Medicine, 101 College St, 1031M, New Haven, CT 06519.

## Abstract

**Objectives:** Transformer models for electronic health records require converting clinical data into token sequences, however standardized tokenization and evaluation frameworks are lacking. We introduce DBToken, an open-source library, and bits-per-row (BPR), a metric for comparing tokenization strategies.

**Materials and Methods:** DBToken accepts Medical Event Data Standard (MEDS)-compatible input and supports multiple text, numeric, and temporal tokenization strategies. BPR extends the bits-per-byte metric used in language models to enable comparison across tokenization strategies.

**Results:** DBToken efficiently tokenized data across configurations. BPR identified the vocabulary size associated with the best clinical outcome performance and localized differences in numeric tokenization performance by token class.

**Discussion:** Optimal tokenization strategies for medical foundation models are a subject of active research. DBToken enables reproducible tokenization experiments, while BPR efficiently screens vocabulary sizes and numeric representations before downstream evaluation.

**Conclusion:** DBToken and the BPR metric provide open-source infrastructure for reproducible EHR tokenization and cross-strategy evaluation.

## 1 BACKGROUND AND SIGNIFICANCE

Transformer-based foundation models for electronic health record (EHR) data have proliferated recently, including CLMBR,[1] ETHOS,[2] CEHR-GPT,[3] EventStreamGPT,[4] TransformEHR,[5] MOTOR,[6] CoMET/Curiosity,[7] and Delphi.[8] Regardless of design, these models must convert heterogeneous clinical records into time-ordered token sequences.

This conversion step is a critical but under-resourced bottleneck. While natural language processing (NLP) uses tools like byte-pair encoding (BPE)[9] and SentencePiece,[10] EHR tokenization must handle categorical codes, continuous variables, ordinal values, and temporal dynamics. MEDS-Torch [11] provides some of these capabilities, but the field lacks a flexible tokenization library for systematic comparison. Recent empirical studies show the importance of tokenization decisions on downstream clinical prediction,[12–14] however these studies largely rely on specialized processing pipelines, motivating the need for a general-purpose tokenization library.

Efficient evaluation of tokenization strategies is challenging because cross-entropy is not directly comparable across different categorical and numeric tokenization strategies. In NLP, metrics such as bits-per-byte and bits-per-character address this problem by normalizing likelihood to a common tokenization-independent unit.[15,16] While these metrics do not replace downstream evaluation, they can serve as a screen to identify top candidates. Structured clinical data currently lack an analogous metric that accommodates the complexities of EHR data.

To address these gaps, we introduce DBToken, an open-source library for mapping EHR data to token sequences, and develop a density-adjusted bits-per-row (BPR) metric for comparison across strategies. Compatible with the Medical Event Data Standard (MEDS),[15] DBToken supports atomic concepts or BPE vocabularies, multiple numeric strategies (discrete quantile bins, continuous distribution scaling, categorical levels), configurable fused or factored layouts, and flexible temporal encoding (age, calendar time, elapsed time). BPR applies bin-width normalization and Jacobian corrections to express likelihoods in the original data space, enabling direct comparison.

## 2 OBJECTIVES

The objectives of this work are:

1. to develop an open-source, configurable tokenization library (DBToken) that provides a unified pipeline for converting tabular EHR data into token sequences across multiple categorical, numeric, and temporal encoding strategies;
2. to introduce a density-adjusted bits-per-row (BPR) evaluation metric that enables information-theoretic comparison between models trained using different tokenization strategies; and
3. to demonstrate these tools by training and comparing GPT-style clinical language models under multiple tokenization configurations on a large-scale EHR dataset, and to provide an interactive dashboard for transparent inspection of how each configuration transforms clinical events into tokens.

## 3 MATERIALS AND METHODS

### 3.1 Tokenization Framework

DBToken is implemented in Python using Polars,[17] NumPy,[18] rustbpe,[19] and tiktoken.[20] It accepts a five-column MEDS-compatible tabular scheme consisting of (`id, time, item, text_value, numeric_value`) that uses a convention to create a conceptual separation between event type (item) and tokenizable non-numeric content (`text_value`), rather than placing all nonnumeric information within the MEDS code field. This formulation allows for event-type dependent tokenization routing and efficient text-based tokenization of the `text_value` field. For example, instead of representing a hemoglobin lab using a delimited `code` such as “`LAB//hemoglobin`”, this observation would be represented as item “`LAB`” and `text_value` “`hemoglobin`”.

The library operates in two stages: a training phase that learns vocabularies, numeric transformation parameters, and temporal boundaries from the training cohort, and an encoding phase that applies these learned parameters to map tabular data to output sequences. Discrete tokenization schemes output a single integer vector of token IDs, and continuous schemes output an additional aligned vector of scaled continuous measurements. The framework is organized around the following four configurable axes.

#### 3.1.1 Text tokenization

Text can be tokenized in concept mode (each unique `text_value` is assigned a unique token) or BPE mode which uses byte-pair encoding to generate a vocabulary across all `text_value` entries. A threshold can be set to automatically route classes exceeding a certain number of unique values to BPE.

#### 3.1.2 Numeric value representation

Numeric values within each (`item`, `text_value`) group are tokenized using one of three methods: (1) Discrete tokenization into configurable quantile bins (Q-tokens), (2) Continuous-value tokenization using user- or automatically-selected, best-fit transformations (normal, lognormal, gamma, min-max) for centering and scaling, or (3) Level-based tokenization for cor (`item`, `text_value`) groups with a small number of unique values (i.e. dosage data with fixed levels).

#### 3.1.3 Numeric value arrangement

In factored mode, numeric values are represented at a separate token position from the associated concept (eg, [`hemoglobin`, `Q3`]). In fused mode, the numeric value is incorporated into the associated concept, producing a single compound token (e.g., `[hemoglobin::Q3]` in discrete fused). Fused placement is restricted to concept-mode classes because BPE does not provide an unambiguous token with which to merge the numeric value.

#### 3.1.4 Temporal encoding

Temporal information is encoded using time deltas, calendar-time milestones, and patient age. Inter-event time deltas can be partitioned into scales where binning or scaling can be separately applied. Calendar-time milestone tokens are emitted when specific temporal boundaries are crossed (e.g. month of the year, or shift change times for hospitalized patients). Patient age is represented via birth-date-derived age tokens emitted on year boundaries.

### 3.2 Bits-Per-Row

In NLP, bits-per-byte (BPB) and bits-per-character (BPC) enable comparison across tokenization strategies by normalizing the model-assigned negative log-likelihood of a text sequence to UTF-8 bytes or characters.[15,16] We adapt this principle to structured clinical data, using rows in the source data table as the common reference unit, and call this concept bits-per-row (BPR).

BPR measures the model-assigned negative log-likelihood of longitudinal clinical data, normalized per row. For categorical tokens, the model-assigned negative log-probability (base-2) is summed across all categorical tokens in the row. For numeric tokens, density corrections are used to convert assigned probabilities to negative log-density in the original measurement units. For discrete tokens, the density is computed by dividing the predicted probability mass assigned to a bin by the width of the bin in the original measurement units. For continuous values, the predicted density in transformed space is converted to density in the original measurement units using the Jacobian of the scaling transformation.[21] BPR can be decomposed into six subcategories (item, text, numeric, time, level, and fused) for a granular assessment of model performance across tokenization axes (Table 1). BPR is computed directly from model-assigned likelihoods allowing for computation during training without significant overhead.

**Table 1.**
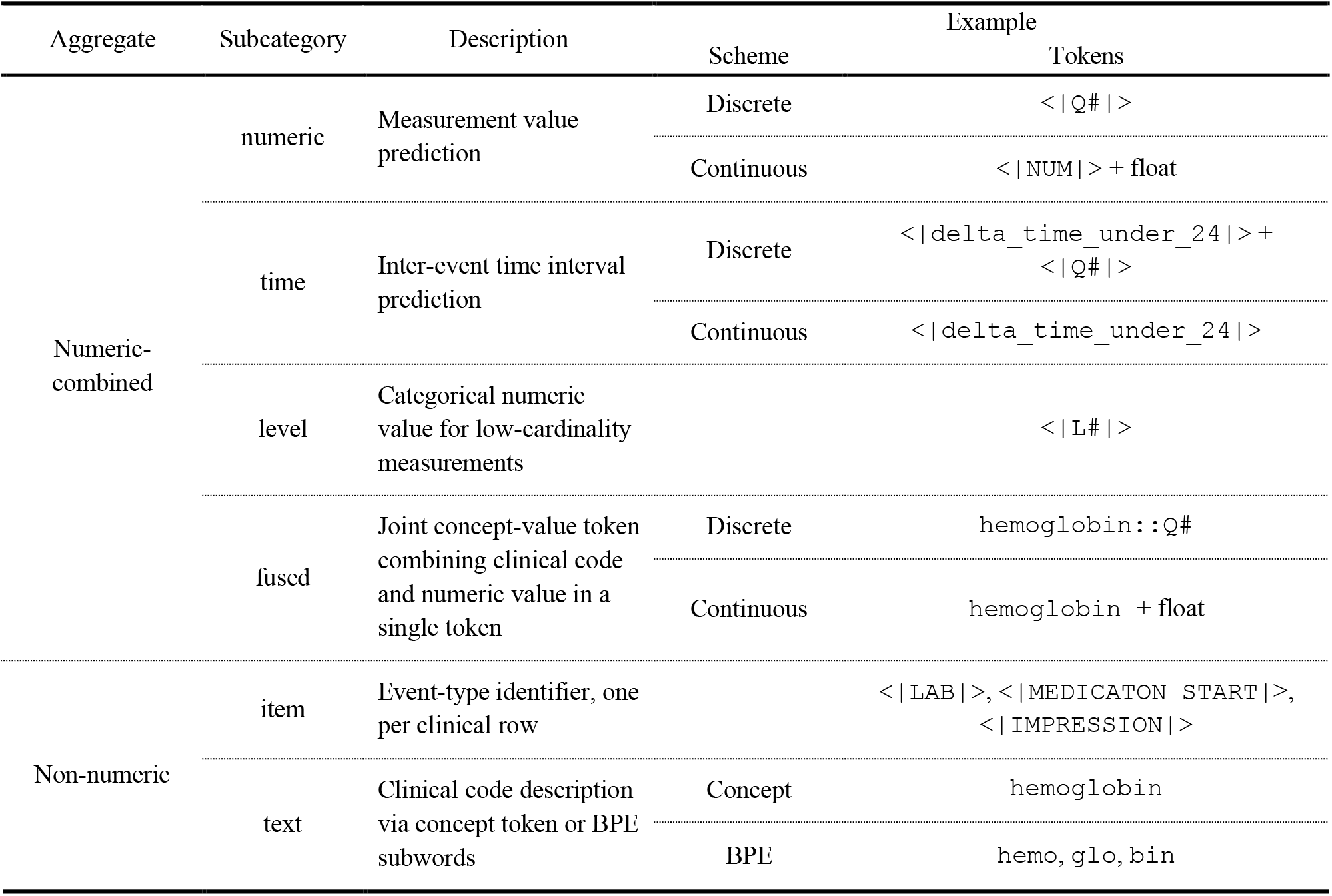
BPR subcategory breakdown.

### 3.3 Data

We used MIMIC-IV version 3.1 [22] and MIMIC-IV-ED version 2.2,[23] a publicly available de-identified EHR dataset containing over 364,000 unique patients. Data were extracted and preprocessed using the MIMIC-IV-MEDS repository [24] and converted to the 5 column format derived from MEDS format.[25] The dataset was split at the patient level into training (80%), validation (10%), and test (10%) sets.

### 3.4 Model Architecture and Training

We evaluated DBToken across two tokenization axes: BPE vocabulary size (2,048 to 131,072) and numeric representation (10 to 100 quantile bins versus continuous with a 16,384 categorical token vocabulary). All configurations used BPE text tokenization for items with more than 20 unique text values, a 24-hour time-delta threshold, and no milestone tokens. The GPT-2 architecture [14] was used for discrete configurations and the related MultivariateGPT [14,19] was used for the continuous configuration. Both models had approximately 110 million parameters, with 12 layers, 768-dimensional embeddings, 12 attention heads, and a 1,024-token context window. Tokenization used one CPU core, and models were trained on one NVIDIA H200 GPU. BPR was computed at each evaluation checkpoint.

### 3.5 Evaluation Procedures

BPR and validation loss were estimated from 3,000 validation batches and 95% confidence intervals were calculated using 1,000 bootstrap resamples. Each vocabulary configuration was evaluated on diagnosis prediction in 2,000 held-out hospital encounters. Models were conditioned on records of the hospital course and were used to generate 64 Monte Carlo simulations of hospital diagnoses. Diagnosis probabilities were estimated from sampling frequencies and compared with diagnoses recorded within 24 hours of discharge using Area Under the Receiver Operator Curve (AUROC).

### 3.6 Interactive Dashboard

To support exploration and communication tokenization strategies, we developed a web-based visualization for the tokenizer which aligns data rows with the tokens emitted and allows users to control tokenization parameters (Figure 1).

**Figure 1.**
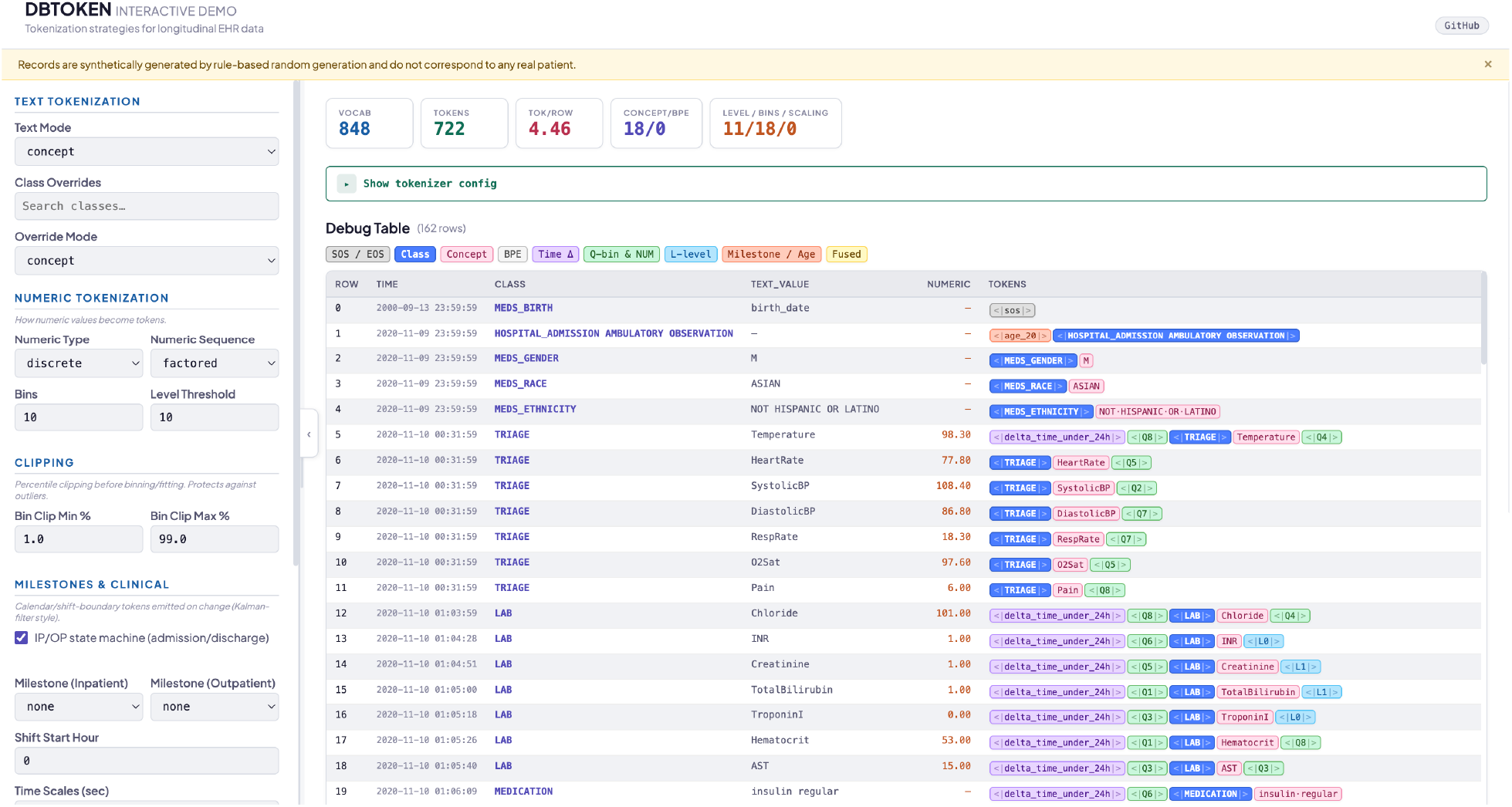
Interactive dashboard view. Left: sidebar with tokenization configuration controls. Center: per-patient token stream with row-level alignment to source events. Right: vocabulary and numeric distribution inspector.

### 3.7 Code Availability

Code is available on github at [https://github.com/AJLozaLab/dbtoken_demo] and on PyPi at [https://pypi.org/project/dbtoken/0.1.0/]. The demonstration is available at [https://dbtoken-demo.lozalab.org/].

## 4 RESULTS

DBToken efficiently tokenized data across configurations. Tokenizing with configurations using BPE and discrete numeric representations was slowest (50,000 rows/second on a single CPU) and configurations with concept-based text tokenization and continuous values were fastest (350,000 rows/second). Details are in **Table S1**.

Table 2 summarizes the tokenization characteristics for five BPE-derived vocabulary sizes. Nearly all compression was reached at a vocabulary size of 16,348. Per-token validation loss increased with vocabulary size (Figure 2A), whereas BPR showed an optimal performance at an estimated vocabulary size of 20,144 (Figure 2B). This optimum vocabulary size estimated from BPR was similar to the estimated optimal vocabulary size estimated from hospital diagnosis prediction of 24,733 (Figure 2C).

**Table 2.** Tokenization configurations and model summary.

| vocab | tokens | tok/seq | p1 | p5 | p10 | p25 | p50 | p75 | p90 | p95 | p99 |
| --- | --- | --- | --- | --- | --- | --- | --- | --- | --- | --- | --- |
| 2k | 1,753,518,957 | 6,010.3 | 7 | 30 | 118 | 299 | 1,159 | 4,592 | 14,085 | 26,329 | 77,118 |
| 4k | 1,561,447,974 | 5,351.9 | 7 | 26 | 108 | 269 | 990 | 3,993 | 12,463 | 23,450 | 70,039 |
| 8k | 1,488,663,711 | 5,102.4 | 7 | 23 | 102 | 251 | 906 | 3,734 | 11,839 | 22,412 | 67,607 |
| 16k | 1,467,609,891 | 5,030.2 | 7 | 22 | 99 | 245 | 877 | 3,647 | 11,658 | 22,134 | 67,060 |
| 32k | 1,463,896,773 | 5,017.4 | 7 | 22 | 98 | 243 | 872 | 3,632 | 11,625 | 22,086 | 66,965 |
| 65k | 1,463,463,935 | 5,016.0 | 7 | 22 | 98 | 243 | 871 | 3,630 | 11,621 | 22,079 | 66,959 |
| 131k | 1,463,090,663 | 5,015.7 | 7 | 22 | 98 | 243 | 871 | 3,629 | 11,620 | 22,078 | 66,957 |

**Figure 2.**
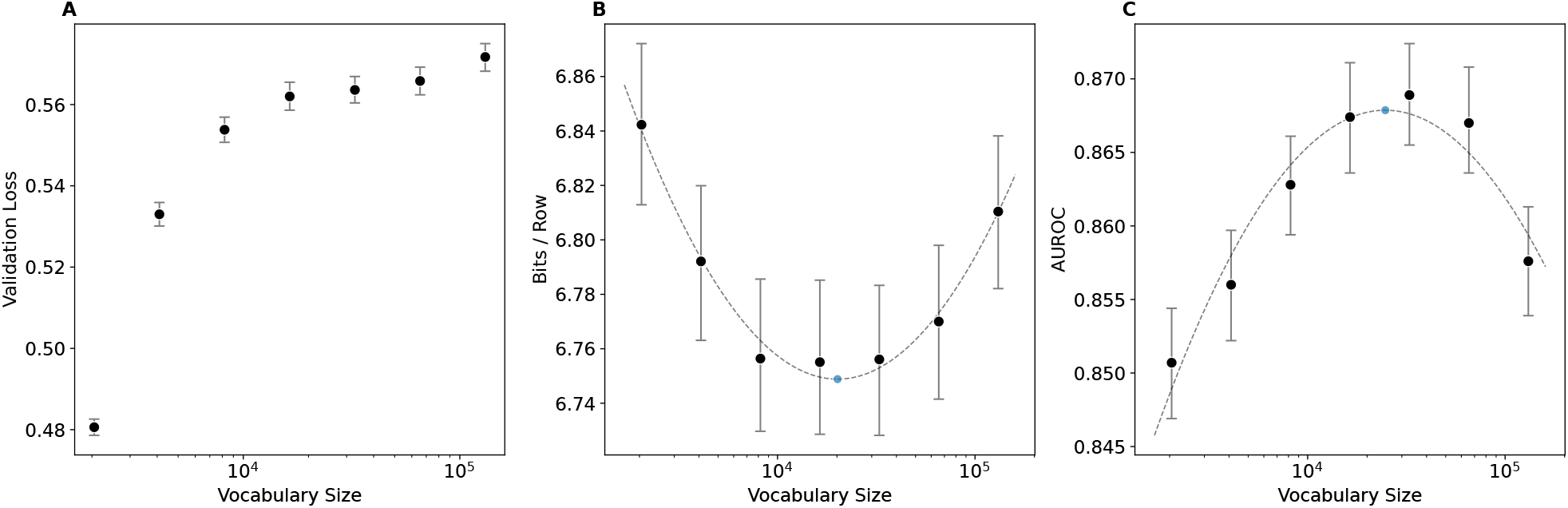
Evaluation metrics across BPE vocabulary sizes for the discrete factored tokenization strategy. (A) Per-token validation loss. (B) Density-adjusted BPR; lower is better (C) Downstream AUROC on ICD code prediction from Monte Carlo sampling; higher is better.

We next compared discrete and continuous numeric tokenization strategies using a 16k vocabulary. Overall, the continuous model achieved lower total BPR (6.631 [95% CI, 6.603–6.660]) than each discrete configurations except the 100-bin model, which achieved a BPR of 6.551 [95% CI, 6.524–6.579] (Figure 3A). For non-time numeric measurements, including laboratory values and vital signs, the continuous model achieved lower BPR than all discrete models (Figure 3B). In contrast, for time-delta tokens, the continuous model had higher BPR than the 40- and 100-bin discrete models (Figure 3C). Inspection of individual time-delta predictions showed that the discrete model’s softmax distribution captured multiple modes across bins, whereas the continuous model’s unimodal output distribution could not capture this complex distribution (Figure S1). BPR contributions from nonnumeric item and text tokens were similar across all configurations (Figure 3D).

**Figure 3.**
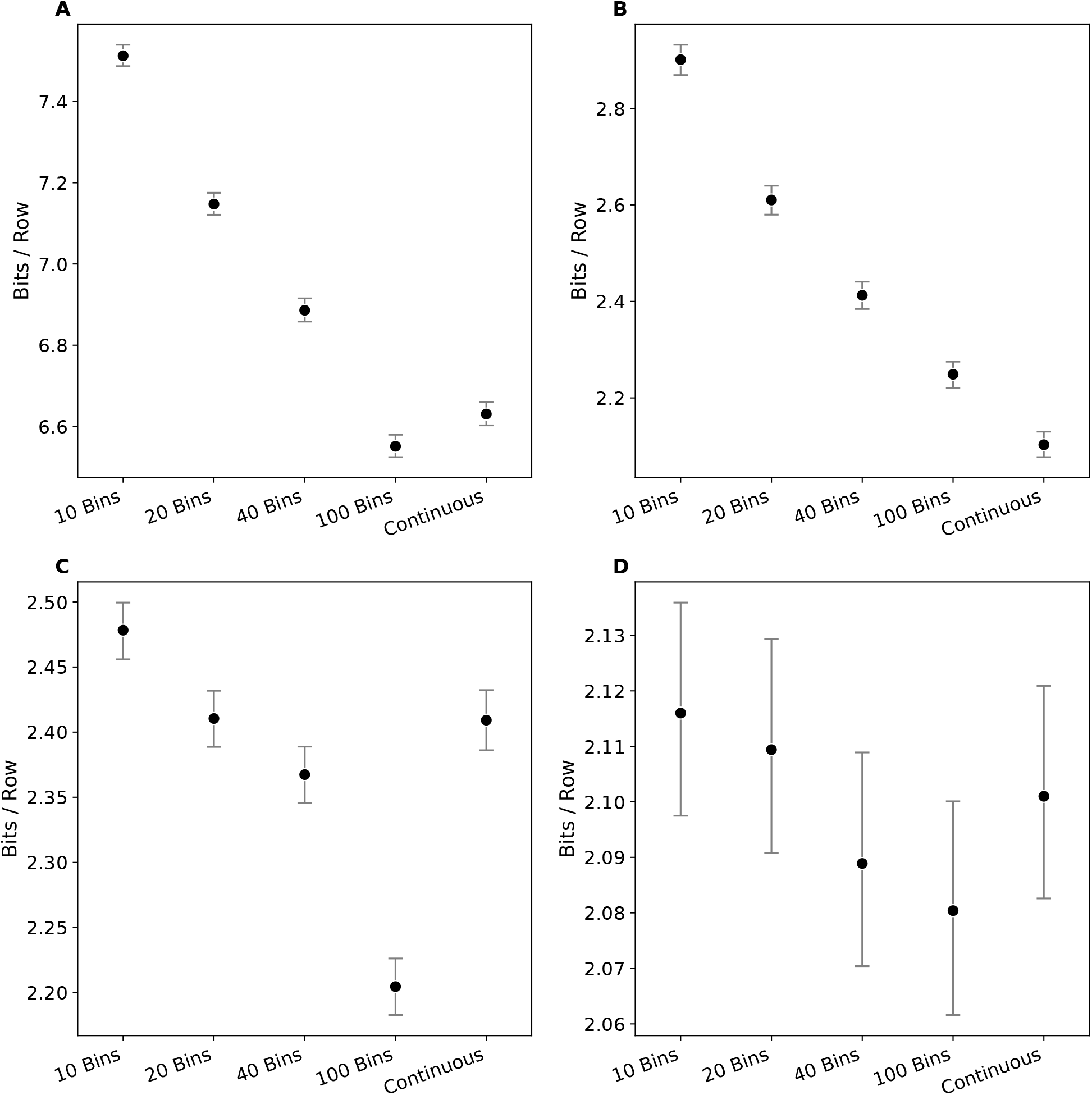
BPR decomposition across discrete bin counts and continuous factored tokenization, all at 16k BPE vocabulary. (A) Total BPR. (B) Non-time numeric BPR (lab values, infusions, vital signs, triage and other clinical measurements). (C) Time-delta BPR. (D) Non-numeric BPR (item and text tokens).

## 5 DISCUSSION

This work introduces DBToken, a configurable tokenization library for EHR data, and bits-per-row (BPR), an information-theoretic metric for comparing tokenization strategies. DBToken efficiently generates token streams from MEDS-compatible data across a range of configurations. The library provides a highly configurable framework encompassing BPE-based text tokenization; discrete, continuous, and level-based numeric representations; fused and factored numeric-value locations; and multiple temporal encoding strategies including time deltas, calendar milestones, and patient age.

BPR enables comparison across tokenization strategies by normalizing model-assigned likelihood to a common unit of data and is computable during training. In a sweep of vocabulary size, next-token loss does not produce a useful ranking for downstream task performance, whereas BPR was correlated with downstream task performance. Decomposition of BPR by token subtype further showed that continuous representations performed well for clinical measurements, while discrete representations better captured multimodal time-delta distributions. This limitation of unimodal time prediction warrants further study because existing clinical sequence models, including Delphi,[8] use similar unimodal formulations.

Several limitations remain. First, validation is currently limited to the MIMIC-IV dataset with 110M parameter models. The results for vocabulary size and numeric bin count may differ across institutions, data sources, and model scales. Second, the library offers four numeric transformations. Third, BPR requires consistent row definitions and measurement space and could limit comparisons across data sets. Finally, while BPR is an efficient metric that can be computed during training, it is intended as a proxy for large-scale tokenization method screening experiments and does not replace the need for downstream clinical evaluation. Further characterization of the correspondence between BPR and a range of downstream clinical tasks is needed and will be the subject of future work.

## 6 CONCLUSION

DBToken provides an open-source framework for configurable tokenization of structured EHR data, and the accompanying density-adjusted bits-per-row (BPR) metric enables principled, information-theoretic comparison across tokenization strategies. Together, these tools support reproducible, computationally efficient screening and selection of candidate tokenization approaches for clinical foundation models.

## Data Availability

MIMIC-IV and MIMIC-IV-ED are available online at PhysioNet (https://physionet.org/content/mimiciv/ and https://physionet.org/content/mimic-iv-ed/) under a credentialed-access Data Use Agreement. Researchers must complete human-subjects research training and agree to the data use restrictions.

https://physionet.org/content/mimic-iv-ed/

## 7 ACKNOWLEDGMENTS

Dr. Loza receives funding through UL1 TR001863, The Hartwell Foundation, and the ARIA foundation. This publication’s contents are solely the responsibility of the authors and do not necessarily represent the official views of NIH or the VA.

