## Supplementary Material for "DBToken: A Database Tokenizer for Medical Event Foundation Models"

### Supplementary Materials

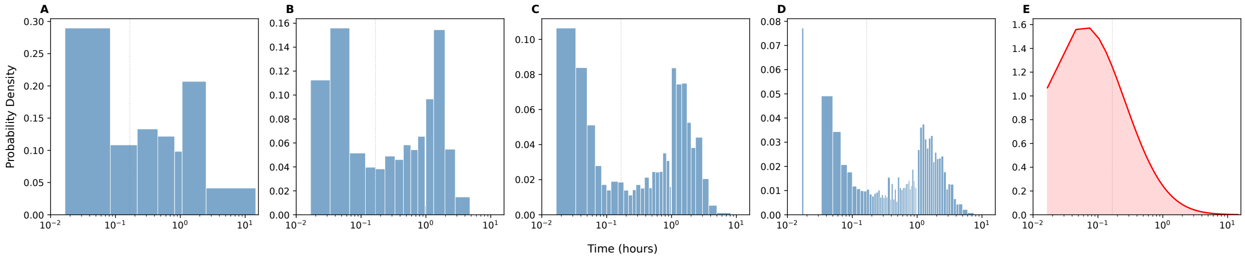


**Supplementary Figure S1.** Predicted time-to-next-event distributions under different numeric tokenization strategies. Panels A–D show predictions using increasing numbers of discrete bins, and panel E shows the prediction using continuous-value tokenization. The discrete formulations capture a multimodal waiting-time distribution that is not represented by the unimodal continuous formulation.

**Supplemental Table 1.** Tokenization speed across configurations

| Text Mode | Bins | Numeric Location | Numeric Type | Vocab Size | Rows Per Second (avg) | Rows Per Second (std) |
|---|---|---|---|---|---|---|
| concept | 10 | factored | discrete | 11536 | 238873 | 3266 |
| concept | | factored | continuous | 11536 | 345099 | 6834 |
| concept | 10 | fused | discrete | 17707 | 242252 | 1869 |
| concept | | fused | continuous | 12361 | 357456 | 2758 |
| concept | 50 | factored | discrete | 11576 | 200230 | 1011 |
| concept | | factored | continuous | 11576 | 352054 | 947 |
| concept | 50 | fused | discrete | 37187 | 195170 | 6444 |
| concept | | fused | continuous | 12401 | 351341 | 4375 |
| concept | 100 | factored | discrete | 11626 | 160012 | 3049 |
| concept | | factored | continuous | 11626 | 329742 | 4081 |
| concept | 100 | fused | discrete | 61537 | 154998 | 3962 |
| concept | | fused | continuous | 12451 | 349460 | 4942 |
| bpe | 10 | factored | discrete | 1024 | 51031 | 1584 |
| bpe | | factored | continuous | 1024 | 55751 | 1500 |
| bpe | 10 | factored | discrete | 2048 | 51800 | 264 |
| bpe | | factored | continuous | 2048 | 53588 | 2393 |
| bpe | 10 | factored | discrete | 4096 | 53068 | 1115 |
| bpe | | factored | continuous | 4096 | 57101 | 518 |
| bpe | 10 | factored | discrete | 8192 | 52351 | 1465 |
| bpe | | factored | continuous | 8192 | 55025 | 1171 |
| bpe | 10 | factored | discrete | 16384 | 52582 | 1020 |
| bpe | | factored | continuous | 16384 | 56094 | 762 |
| bpe | 10 | factored | discrete | 17383 | 53010 | 758 |
| bpe | | factored | continuous | 17383 | 56728 | 496 |
| bpe | 10 | factored | discrete | 17383 | 53392 | 315 |
| bpe | | factored | continuous | 17383 | 57274 | 990 |
| bpe | 10 | factored | discrete | 17383 | 53952 | 50 |
| bpe | | factored | continuous | 17383 | 58175 | 393 |
